# Neuroimaging Inequities in Africa: Health system readiness, diagnostic capacity, and an embedded case study of portable ultra-low-field MRI implementation in Ethiopia

**DOI:** 10.64898/2026.10.23.26366362

**Authors:** Gebeyehu Begashaw Abate, Petter Clemensson, Niall J. Bourke, Carly Bennallick, Steve Williams, Ricardo Twumasi, Vincent Giampietro

**Affiliations:** Department of Neuroimaging, Institute of Psychiatry, Psychology & Neuroscience, King’s College London, UK; Department of Psychosis Studies, Institute of Psychiatry, Psychology & Neuroscience, King’s College London, UK; University of Gondar, Ethiopia

**Keywords:** ultra-low field MRI, neuroimaging, global health equity, Africa, LMICs

## Abstract

Neurological conditions in Africa are an increasingly significant public health burden. Conditions such as stroke, dementia, and central nervous system infections impose a disproportionate burden in resource constrained regions, highlighting the need to strengthen diagnostic capacity. Magnetic Resonance Imaging (MRI) and computed tomography (CT) are essential technologies for the diagnosis and management of these conditions. However, the availability and affordability of these advanced diagnostic technologies remain limited across the continent. A low-cost, portable ULF-MRI system may help expand access to neuroimaging services across diverse settings by overcoming some of the infrastructural and affordability challenges. The aim of this study was to assess the diagnostic imaging capacity and health system readiness for advanced neurological services in Africa and to examine the feasibility of integrating portable ULF-MRI into routine clinical care in a low-resource setting, specifically in Ethiopia. Quantitative data on MRI and CT capacity and selected health system infrastructure indicators were obtained from the World Health Organization Global Health Observatory Database. The qualitative case study was based on a recently deployed 0.064T ULF-MRI Hyperfine Swoop® system at a regional specialized hospital in Ethiopia for clinical use. The results showed substantial gaps in diagnostic imaging capacity, workforce availability, and supporting infrastructure across Africa. MRI and CT density in Africa stood at 0.11 MRI and 0.42 CT scanners per million population, respectively. The Ethiopian case study suggested that ULF-MRI may be a promising approach to expanding neuroimaging access because it can be integrated into routine clinical practice with relatively modest infrastructure and personnel requirements. These findings underscore major diagnostic inequities across African health systems and suggest that portable ULF-MRI may offer a feasible approach to expanding access to neuroimaging services in resource-constrained settings.

## Introduction

Neurological conditions in low-and middle-income countries (LMICs) have become an increasingly significant public health burden. Conditions, such as stroke, dementia, central nervous system infections, cancer, and epilepsy, impose a disproportionate burden in this region, highlighting the need for a greater focus on expanding neurological services (1,2). Neuroimaging technologies, such as magnetic resonance imaging (MRI) and computed tomography (CT), play significant roles in the prevention, diagnosis, and interventions of neurological health challenges through characterising (abnormal) structural, functional, and metabolic brain patterns (3,4). Yet, access to such advanced imaging technologies in LMICs is very limited and remains a key barrier to global brain health equity.

Operating advanced neuroimaging technologies like MRI requires an extensive and consistent power supply, trained personnel, and dedicated infrastructure, including a radiofrequency-shielded room (5,6). However, such health system infrastructures are limited in low-resource settings, affecting the availability and accessibility of MRI and CT (7,8). Moreover, the acquisition of such diagnostic technologies is expensive. For example, a conventional MRI scanner costs between one and half million and four million dollars with ongoing additional expenses for maintenance and hardware replacements (9–11). Furthermore, the immobility of MRI scanners can limit accessibility, as the magnet alone weighs between 3000 and 6000 kilograms (10,12).

To address the challenges of affordability, accessibility, and complex infrastructural requirements associated with conventional diagnostic technologies, a new generation of portable Ultra-Low-Field MRI (ULF-MRI) systems (<0.1T compared to 1.5T or 3T for high-field MRI (HF-MRI)) is now emerging as a compelling alternative. These scanners offer the potential to bring neuroimaging procedures to the point-of-care in emergency units, intensive care units (ICU), and remote hospitals (7) through bypassing the logistical and economic constraints of conventional 1.5 or 3T systems. This is particularly relevant for LMICs where neurological conditions are on the rise due to a high rate of infectious diseases, malnutrition, and other socio-economic factors and within the context of a broader neuroimaging ecosystem gap (13,14).

Portable ULF-MRI systems differ significantly from HF-MRI devices. They rely, for example, on the use of permanent magnets instead of high-powered superconducting electromagnets, and, as such, have significantly reduced costs associated to power, cooling, and maintenance. This leads to ULF-MRI systems to be smaller, more mobile, and with substantially reduced need for stringent safety measures and electromagnetic shielding. Additionally, anyone can operate a ULF-MRI scanner after only brief training and local biomedical engineers can perform basic maintenance after minimal preventative training (15,16). Furthermore, health facilities do not need to change their infrastructure to integrate this technology into their neuroimaging workflow. For example, ULF-MRI systems do not need to be connected to a special power grid, as they can run from a normal wall outlet.

Portable ULF-MRI can acquire key imaging sequences including, T1-weighted (T1W), T2-weighted (T2W), fluid-attenuated inversion recovery (FLAIR), and diffusion-weighted imaging (DWI) sequences. These have been used in several studies that reported the clinical applications of ULF-MRI in diverse disease conditions such as stroke (8,16), multiple sclerosis (17,18), dementia (19), demyelinating diseases (20), and epilepsy (21,22). ULF-MRI has also proven effective as a prognostic tool for evaluating optic nerve decompression after endoscopic endonasal surgery (23) and for detecting hypoxic ischemic brain injury (HIBI) after cardiac arrest (24). Beyond its clinical applications, ULF-MRI also expands the frontiers of neuroscience and/or brain health research through reaching underrepresented populations including communities in low-resource settings, paediatrics, and critically ill patients.

In view of the transformational potential of ULF-MRI, it is essential to note some of the technical limitations of this system. For example, studies have mentioned that ULF-MRI has a low signal-to noise ratio (SNR) compared to HF-MRI due to its lower magnetic field. This compromises the quality of images because they are of lower resolution, which could lead to clinicians missing subtle diagnostic details and brain lesions (3,4,25). Coarse spatial resolution (∼2-3 mm), long scan times, and lack of tissue contrast are also issues that need to be addressed (1). However, advancements in hardware (such as improved magnet and coil designs) and software (e.g. deep learning reconstruction and super-resolution of images) may help address some of these limitations (17,26).

Researchers and advocacy groups have suggested frameworks to help pave the way for making MRI accessible and affordable in low resource settings, particularly in sub-Saharan Africa (SSA) (1,27,28). One of the recommendations from these frameworks is to accelerate the adoption of low-cost low-field MRI systems and leverage technological advances such as artificial intelligence and open-source software for further innovations. Accordingly, the Ultra-low-field Neuroimaging In The Young (UNITY) initiative (9), a global collaborative effort, is currently working to implement ULF-MRI scanners (Hyperfine’s Swoop® Portable MRI System) at over 30 clinical and research sites across the world including at 11 research sites in Africa.

This research aims to address key evidence gaps regarding neuroimaging capacity in Africa. Firstly, it provides empirical evidence of global health inequities in neuroimaging services through a comprehensive, longitudinal, and region-wide analysis of MRI and CT density. Secondly, although ULF-MRI has been proposed as a potential solution to addressing accessibility and affordability challenges associated with neuroimaging in Africa, there is limited evidence regarding the adoption and integration of this technology in real-world African healthcare systems.

This study therefore evaluates the implementation of ULF-MRI in the Ethiopian healthcare system through a qualitative case study and provides insight into its feasibility and integration within routine clinical practice. Finally, linking diagnostic technologies with health system infrastructure in Africa is essential to develop viable and innovative solutions for strengthening neuroimaging capacity in the region. Accordingly, this study analyses the existing infrastructure, including the healthcare workforce and electrification of healthcare centres, both of which are essential prerequisite for the delivery of advanced imaging services.

## Methods and materials

### Study design

This study utilized a multi-method research design, combining descriptive quantitative analysis of diagnostic imaging availability and health system infrastructure in Africa with an embedded case study examining the integration of ULF-MRI in a low-resource healthcare setting. Ethical approval for this study was obtained from King’s College London (LRS/RGO-24/25-47006).

### Material

The Hyperfine Swoop® system is a highly mobile (motorised), cart-based MRI head imager, designed to roll through standard doorways and elevators and to park at the bedside in wards, and intensive care units (Figure 1). Its compact, self-shielded design allows safe deployment in routine clinical spaces without a built-in MRI suite. The scanner’s head coil is made of a solid, clear plastic designed to keep the subject’s head stable during the scan and can be easily disinfected between subjects. It also contains transmitters and receivers to capture signals to generate the images. Padding is usually placed either side of the head to keep the subjects head still during scanning and minimise motion. The coil is surrounded by a perforated scanning cage with multiple access doors that allow for positioning (Rowand et al., 2025).

**Figure 1.**
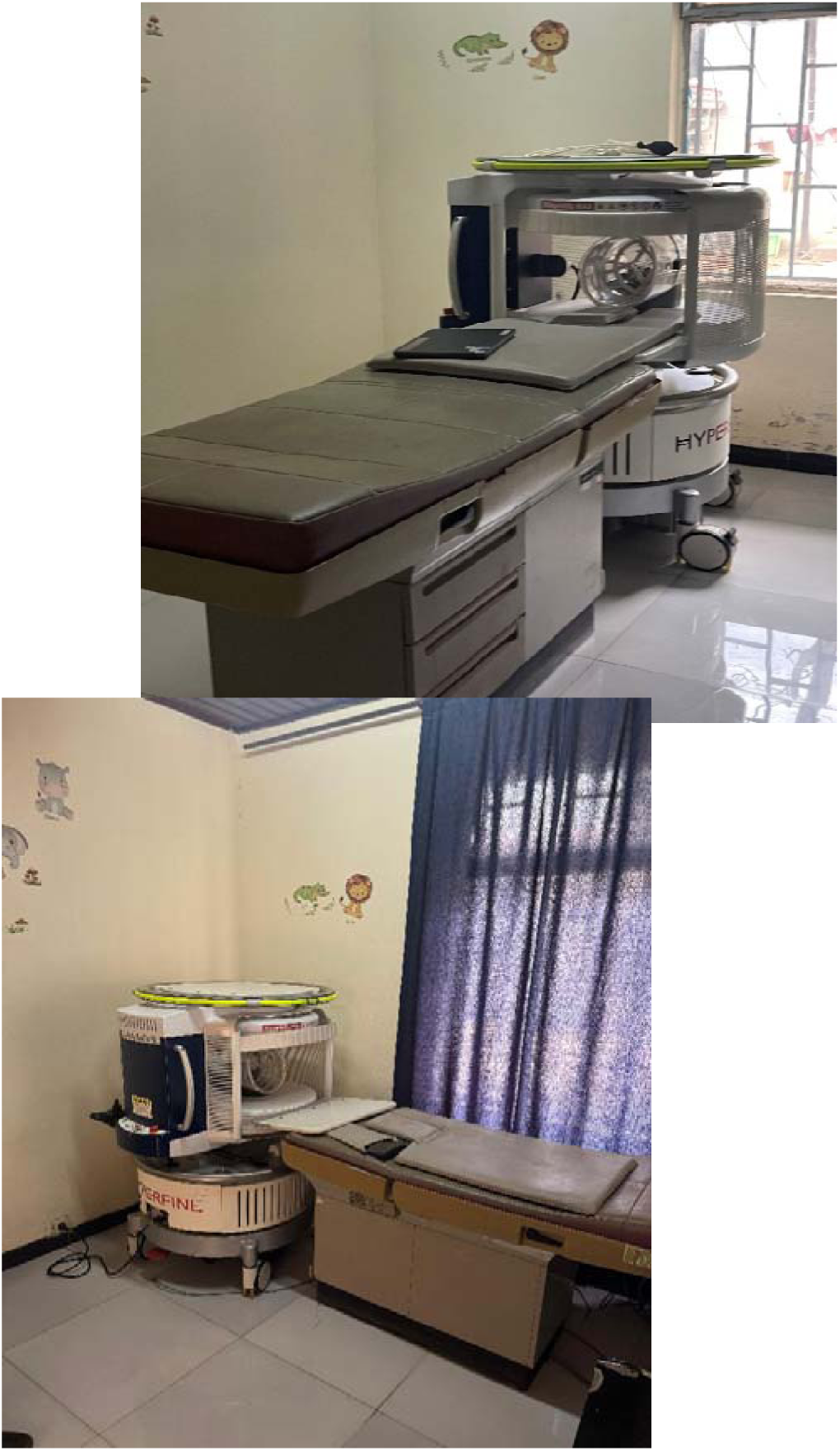
Swoop® Portable MR Imaging® System at Felege Hiwot Hospital, Ethiopia.

The system is operated via an iPad-based interface (29). Crucially, it is built around a permanent magnet (0.064 Tesla), avoiding the superconducting electromagnets and cryogenic cooling required by conventional MRI scanners (see Table 1 for more specifications). The mobility of the scanner is powered by an internal battery that is charged while being plugged in via the power cord. The scanner used in the case study was deployed in Ethiopia in 2022, initially for research purpose, and over 300 infants’ brain were scanned over a two-year period.

**Table 1.**
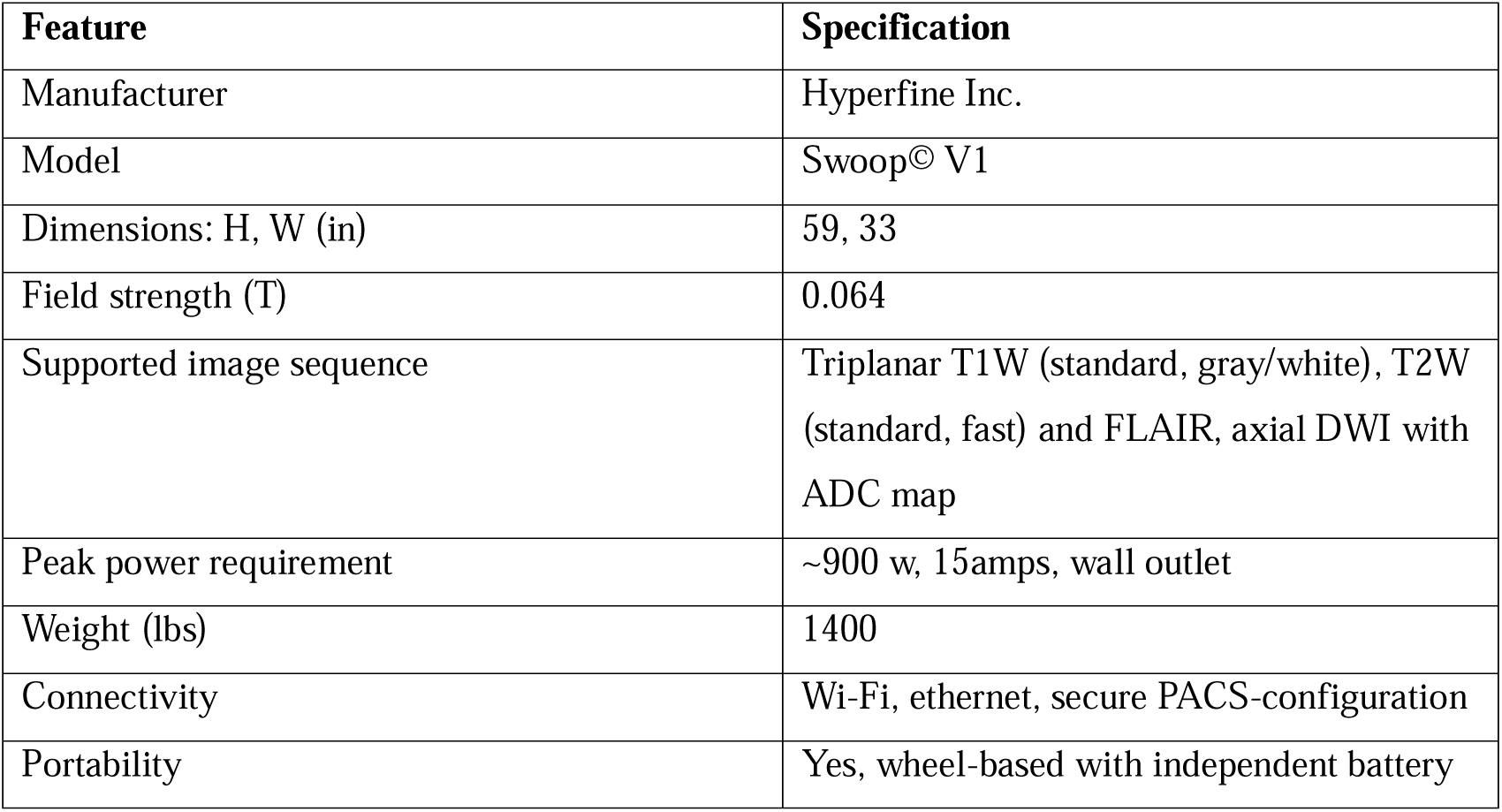
Characteristics of the ULF-MRI at Felege Hiwot Hospital, Ethiopia.

| **Feature** | **Specification** |
| --- | --- |
| Manufacturer | Hyperfine Inc. |
| Model | Swoop© V1 |
| Dimensions: H, W (in) | 59, 33 |
| Field strength (T) | 0.064 |
| Supported image sequence | Triplanar T1W (standard, gray/white), T2W (standard, fast) and FLAIR, axial DWI with ADC map |
| Peak power requirement | ~900 w, 15amps, wall outlet |
| Weight (lbs) | 1400 |
| Connectivity | Wi-Fi, ethernet, secure PACS-configuration |
| Portability | Yes, wheel-based with independent battery |

### Study setting and participants

The case study component of this research was conducted at Felege Hiwot comprehensive specialized hospital in northwestern Ethiopia, which is one of the main hospitals in Bahir Dar city, located in Amhara Regional State. Felege Hiwot Hospital primarily offers imaging services using CT and X-ray modalities and has recently added the Hyperfine Swoop® portable MRI system to its imaging services. It is the first hospital in Ethiopia to deploy the ULF-MRI for clinical use. However, the hospital does not have a high-field MRI system. The study participants included radiologists, radiology technologists, biomedical engineers, and nurses working at the hospital. They were selected purposively based on their direct experience with and involvement in the implementation of ULF-MRI.

### Data collection procedures and analysis

Quantitative data on MRI and CT scanners, healthcare infrastructure indicators (e.g. healthcare facility access to electricity), and health workforce (e.g. physician and biomedical engineers) were collected from the World Health Organization Global Health Observatory database (https://www.who.int/data/gho).These data include the number and density of MRI and CT per million population (full report is available for only 38 African countries), access to electricity in healthcare facilities, and health workforce density per population. The WHO baseline survey was conducted in 2010, with subsequent updates in 2022. WHO initially planned periodic updates (every five years) of the global atlas of medical devices but shifted from fixed reference year surveys to compilation of the latest available country information. The 2022 data then mean the most recent reported data by each country between 2013 and 2021. Additionally, we used the latest dataset (2022) to get data from the Organization for Economic Co-operation and Development (OECD) countries for comparison purposes.

The qualitative data were collected using a semi-structured interview guide. The interviews focused on healthcare professionals experiences operating the ULF-MRI, the challenges encountered during its implementation, and their perspectives on image quality, technology usability, and workflow integration. In addition, the researcher conducted observations at the hospital, with a particular focus on the infrastructure requirements, the feasibility, and usability of ULF-MRI in a real clinical setting. Two site visits were conducted on May 1, 2025, and August 15, 2025. During these visits, the process of integrating the portable MRI system into the existing imaging infrastructure was observed. The quantitative data were analysed using descriptive statistics such as medians and percentages, in R statistical software. Thematic analysis was used to analyse the qualitative data.

## Results

### Density of MRI and CT

The results showed that the density of MRI and CT scanners in Africa is low. In 2022, the median MRI density across 38 African countries was 0.115 MRI scanner per million population (interquartile range [IQR]: 0.00-0.28). In addition, the median CT scanner density was 0.42 (IQR: 0.20-1.15) per million population. Only three of the 38 countries reported an MRI scanner density greater than one per million population, while eight countries reported a CT scanner density greater than one scanner per million population (see Table 2). The analysis also showed that, in 2022, at least 12 African countries had no MRI scanners, and three countries had neither MRI nor CT scanners.

**Table 2:** MRI and CT scan number of units and density for African and OECD countries.

| Country | MRI-units & density/million population (2010) | MRI-units & density/million population (2022) | CT-density 2010/million population | CT density 2020/million population | OECD country | MRI units & density/million population (2022) | CT density 2020/per million population |
| --- | --- | --- | --- | --- | --- | --- | --- |
| Angola | 1(0.05) | 1 (0.05) | 0.48 | 0.42 | Australia | 375(14.71) | 69.9 |
| Benin | 0 (0) | 0 (0) | 0.33 | 0.29 | Austria | 208 (23.1) | 28.3 |
| Botswana | 1(0.5) | 1 (0.49) | 1.025 | 0.99 | Belgium | 133 (11.48) | 23.47 |
| Burkina Faso | 0 (0) | 2 (0.1) | 0.19 | 0.72 | Canada | 389 (10.31) | 14.8 |
| Burundi | 0 (0) | 0 (0) | 0.24 | 0.2 | Chile | 226 (11.82) | 23.3 |
| Cote d'Ivoire | 3 (0.14) | 3 (0.15) | 0.66 | 0.69 | Colombia | 12 (0.24) | 5.9 |
| Cabo verde | 0 (0) | 3 (5.4) | 1.97 | 14.39 | Costa Rica | 4 (0.82) | 5.13 |
| Cameron | 1 (0.05) | 1 (0.04) | 0.71 | 0.63 | Czech Republic | 110 (10.27) | 16 |
| Central Africa | 0 (0) | 0 (0) | 0 | 0 | Denmark | 77 (13.7) | 40.8 |
| Chad | 0 (0) | 1 (0.06) | 0.089 | 0.18 | Estonia | 18 (13.56) | 18.8 |
| Comoros | 0 (0) | 0 (0) | 1.47 | 1.15 | Finland | 159 (28.7) | 16.4 |
| Democratic Republic of the Congo | 0 (0) | 0 (0) | 0 | 0.074 | France | 1034 (15.84) | 18.7 |
| Eritrea | 1(0.19) | 1(0.16) | 0.394 | 0.32 | Germany | 2869 (34.24) | 34.7 |
| Eswatini | 0 (0) | 0 (0) | 2.53 | 2.4 | Greece | 315 (30.22) | 41.8 |
| Ethiopia | 3 (0.04) | 18 (0.16) | 0.24 | 0.42 | Hungary | 48 (4.97) | 9.5 |
| Gabon | 2 (1.4) | 2 (1.2) | 4.06 | 3.59 | Iceland | 7 (20.53) | 49.9 |
| Gambia | 1 (0.58) | 1(0.54) | 0.54 | 1.24 | Ireland | 78 (15.8) | 21.2 |
| Ghana | 2 (0.08) | 7 (0.23) | 0.23 | 0.35 | Israel | 46 (5.31) | 10.1 |
| Guinea | 0 (0) | 0 (0) | 0 | 0 | Italy | 1736 (28.71) | 35.1 |
| Guinea-Bissau | 0 (0) | 0 (0) | 0 | 0 | Japan | 6996 (55.31) | 111.7 |
| Kenya | 7 (0.18) | 7 (0.16) | 0.276 | 0.25 | Korea (South Korea) | 1553(30.29) | 38.9 |
| Madagascar | 0 (0) | 0 (0) | 0.152 | 0.13 | Latvia | 26 (13.79) | 39.2 |
| Malawi | 1(0.07) | 1 (0.06) | 0.34 | 0.30 | Lithuania | 40 (14.7) | 26.8 |
| Mali | 0 (0) | 0 (0) | 0.23 | 0.20 | Luxembourg | 9 (14.38) | 22.4 |
| Mauritania | 3 (0.9) | 3 (0.77) | 1.82 | 1.54 | Mexico | 316 (2.45) | 6.2 |
| Mauritius | 6 (4.65) | 6 (4.82) | 6.2 | 6.43 | Netherlands | 203 (11.85) | 12.0 |
| Namibia | 2 (0.92) | 2 (0.86) | 4.82 | 4.77 | New Zealand | 76 (15.76) | 15.8 |
| Niger | 0 (0) | 0 (0) | 0.196 | 0.17 | Norway | 169 (31.16) * | 28.2 |
| Senegal | 2 (0.16) | 4 (0.24) | 0.398 | 1.19 | Poland | 350 (9.5) | 17.3 |
| Seychelles | 1(11.9) | 1 (10.8) | 11.86 | 10.77 | Portugal | 105 (9.9) | 27.4 |
| Sierra Leone | 0 (0) | 0 (0) | 0.35 | 0.33 | Slovakia | 52 (9.52) | 18.3 |
| South Africa | 12 (0.23) | 12 (0.2) | 1.01 | 0.86 | Slovenia | 26 (12.51) | 18.2 |
| Sudan | 12 (0.28) | 12 (0.32) | 0.8 | 1.13 | Spain | 805 (17.22) | 19.1 |
| Togo | 1 (0.17) | 1(0.15) | 0.83 | 0.73 | Sweden | 191 (18.2) * | 23.7 |
| Uganda | 1(0.03) | 6 (0.13) | 0.33 | 0.44 | Switzerland | 242 (27.50) * | 40.0 |
| Tanzania | 2 (0.04) | 4 (0.07) | 0.14 | 0.17 | Türkiye | 915 (10.85) | 14.4 |
| Zambia | 1(0.08) | 1 (0.07) | 0.23 | 0.21 | United Kingdom | 581 (8.60) * | 9.5 |
| Zimbabwe | 4 (0.32) | 4 (0.28) | 0.48 | 0.42 | United States | 13275 (40.11) | 44.6 |

*Data source: Global health observatory database \*Data sourced from OECD health equipment statistics;*

The disparity is even more evident when comparing African countries with OECD countries. For example, most OECD countries have more than 10 MRI scanners per million population, with a median density of 14.09 (IQR: 10.27 - 23.10). In 2022, a combined total of 105 MRI scanners were available across the 38 African countries, fewer than the number available in many individual OECD countries. For example, Japan (N = 6,996), South Korea (N =1,553), Italy (N = 1,736), Germany (N = 2,869), and the USA (N = 13,275), each have thousands of MRI scanners. A similar disparity is observed in CT scanner density, with a median of 0.42 per million population (IQR: 0.20-1.15) in Africa compared with 22.85 scanners per million population (IQR: 16.30-38.98) in OECD countries. Overall, the analysis showed the median density of MRI scanners in OECD countries was approximately 120 times higher than that observed in Africa, while the median CT density was approximately 54 times higher, underscoring substantial disparities in access to advanced diagnostic imaging technologies.

### Trend analysis of MRI and CT density

Using the 2010 baseline and the latest MRI and CT scanners data in 38 African countries (see Table 2), a trend analysis was conducted to determine whether there have been improvements in the availability of these technologies. The analysis revealed that there was modest growth in MRI units between 2010 and 2022, where it increased from a combined total of 70 to 105 units, representing an increase of approximately 50%. This growth translates into an increase of the median density of MRI scanners from 0.05 per million population to 0.12 per million population. Similarly, a small increase in median CT density per million population was observed from 2010-2020. The median density rose from 0.37 in 2010 to 0.42 per million population in 2020 (see Figure 2). Out of the 15 countries that reported no MRI scanners unit in 2010, 12 of them continued to have no MRI scanners in 2022.

**Figure 2:**
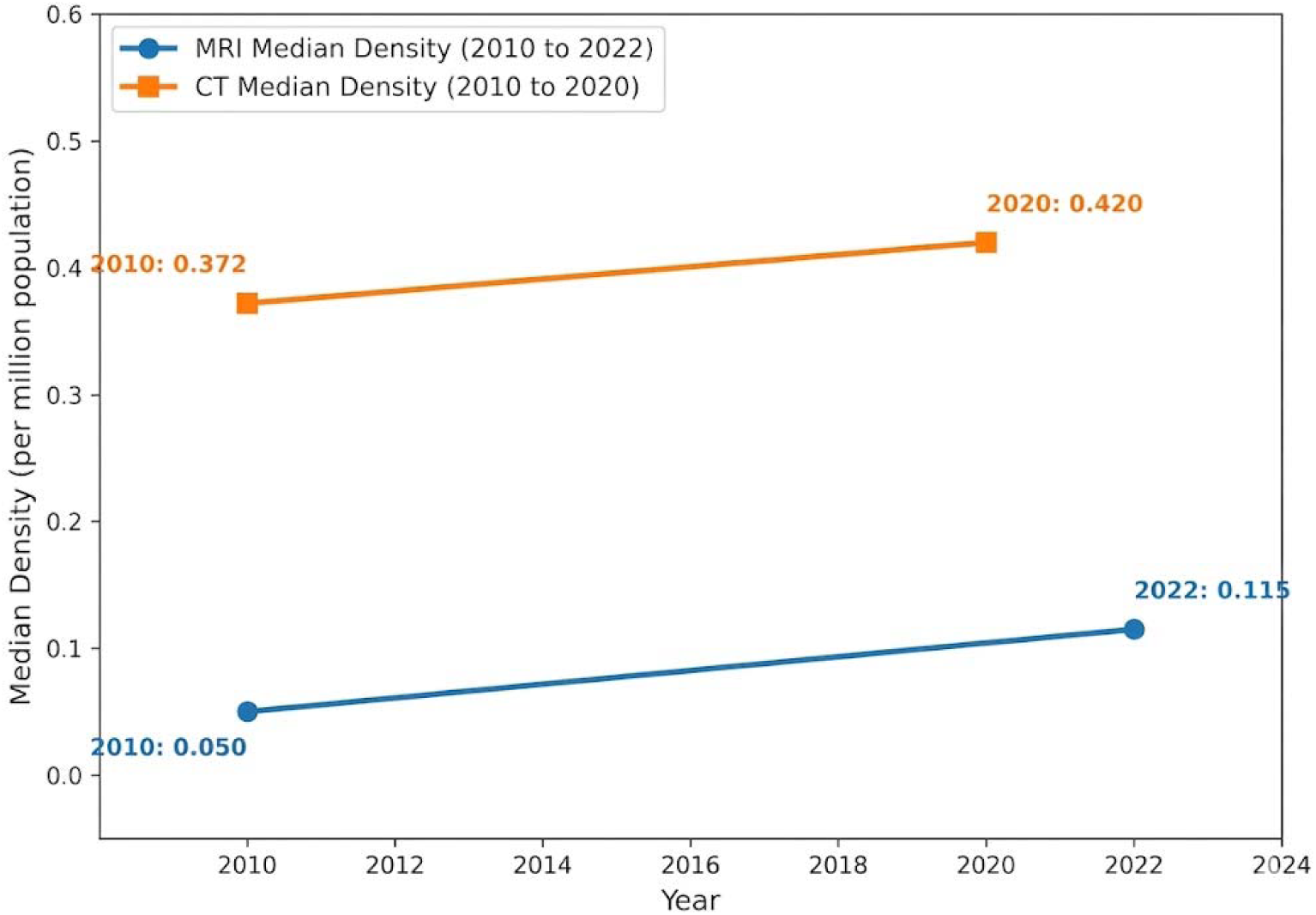
Trends of median density of MRI and CT scans in 38 African countries.

The modest increase in MRI and CT may indicate the gradual expansion of neuroimaging services. However, MRI and CT availability remains extremely low given the population size of Africa and compared with those OECD countries. It is also important to note that most changes were coming from a few countries that added more MRI and CT scanners including Ethiopia, Ghana, and Uganda, while no changes in the absolute number of MRI and CT units were observed in more than two-thirds of the countries.

### Health system infrastructure

This study reveals that there is inadequate health system infrastructure in Africa, particularly in indicators such as access to stable electricity and trained health workforce that are imperative to operate advanced diagnostic technologies like MRI and CT. For example, the median density of physicians per 10,000 population was only 1.9 across the African countries, while the median density for OECD countries was 38 physicians per 10,000 population (see Table 3). Similarly, most African countries have very limited number of biomedical engineers. For example, among countries with available data, most reported fewer than 0.05 biomedical engineers per 10,000 population. In contrast, OECD countries had a median density of approximately 0.41 biomedical engineers per 10,000 population, indicating greater availability of technical personnel to support advanced diagnostic technologies.

**Table 3:** Health workforce (physicians and biomedical engineers) in Africa and OECD countries.

| Country | Physician density/10,000 population | Bio-medical engineers/10,000 population | OECD country | Physician density/10,000 population | Bio-medical engineers/10,000 population |
| --- | --- | --- | --- | --- | --- |
| Angola | 2.02 | No data | Australia | 39.8 | 0.13 |
| Benin | 2.14 | <0.05 (n=16) | Austria | 55 | 0.91 |
| Botswana | 4.28 | 0.23 (n= 52) | Belgium | 33.9 | 0.44 |
| Burkina Faso | 1.07 | <0.05 (n=4) | Canada | 28 | 0.18 |
| Burundi | 0.83 | <0.05 (n= 1) | Chile | 33 | 0.36 |
| Cote d'Ivoire | 2.55 | <0.05 (n= 50) | Colombia | 25 | 0.41 |
| Cabo Verde | 7.3 | No data | Costa Rica | 27 | 0.17 |
| Cameroon | 0.88 | <0.05 (n= 19) | Czech Republic | 44 | <0.05 |
| Central African Republic | 1 | No data | Denmark | 45 | 0.79 |
| Chad | 0.8 | <0.05 (12) | Estonia | 35 | 0.46 |
| Comoros | 4.24 | No data | Finland | 36 | 1.55 |
| Congo | 4.98 | <0.05 (n= 1) | France | 33 | 0.09 |
| Democratic Republic of the Congo | 2.08 | <0.05 (n = 100) | Germany | 45 | 0.25 |
| Equatorial Guinea | 1.53 | No data | Greece | 66 | 0.28 |
| Eritrea | 0.89 | No data | Hungary | 35 | 0.41 |
| Eswatini | 5.57 | <0.05 (n= 1) | Iceland | 44 | 1.68 |
| Ethiopia | 1.56 | 0.05 (n= 500) | Ireland | 39 | 0.69 |
| Gabon | 5.21 | No data | Israel | 38 | 2.34 |
| Gambia | 0.9 | <0.05 (n = 3) | Italy | 42 | 1.4 |
| Ghana | 2.89 | 0.16 (n =450) | Japan | 26 | 3.23 |
| Guinea | 1.15 | 0 | Korea (South Korea) | 26 | <0.05 |
| Guinea-Bissau | 2.49 | No data | Latvia | 34 | 1.12 |
| Kenya | 2.6 | <0.05 (n = 50) | Lithuania | 61 | 0.87 |
| Lesotho | 1.84 | No data | Luxembourg | 30 | N/A |
| Liberia | 1.78 | 0 | Mexico | 26 | 0.29 |
| Madagascar | 1.03 | No data | Netherlands | 39 | 0.29 |
| Malawi | 0.84 | <0.05 (n =5) | New Zealand | 35.7 | 0.06 |
| Mali | 3.17 | No data | Norway | 50 | <0.05 |
| Mauritania | 2.61 | No data | Poland | 40 | <0.05 |
| Mauritius | 30.15 | No data | Portugal | 59 | 2 |
| Mozambique | 0.91 | <0.05 (n= 1) | Slovakia | 37 | 0.12 |
| Namibia | 5.58 | <0.05 (n= 1) | Slovenia | 34 | 0.84 |
| Niger | 0.38 | No data | Spain | 43 | 0.22 |
| Nigeria | 4.14 | <0.05 (n= 280) | Sweden | 44 | 1.15 |
| Rwanda | 1.77 | <0.05 (n =15) | Switzerland | 45 | 0.14 |
| Sao Tome and Principe | 4.64 | No data | Türkiye | 22 | 0.12 |
| Senegal | 1.24 | <0.05 (n =23) | United Kingdom | 33 | 0.53 |
| Seychelles | 16.33 | 0.21 (n =2) | United States | 37 | 0.68 |
| Sierra Leone | 1.34 | <0.05 (n= 1) |  |  |  |
| South Africa | 7.66 | 0.05 (n= 300) |  |  |  |
| South Sudan | 1.49 | No data |  |  |  |
| Togo | 0.59 | No data |  |  |  |
| Uganda | 2.06 | <0.05 (n= 49) |  |  |  |
| United Republic of Tanzania | 1.75 | <0.05 (n= 20) |  |  |  |
| Zambia | 1.63 | <0.05 (n =18) |  |  |  |
| Zimbabwe | 1.86 | No data |  |  |  |

*Data source: Global Health Observatory Database*

In this study, we also analysed access to electricity in healthcare facilities for over a dozen African countries for which data were available (see Table 4). The results show that in some countries, nearly half of the healthcare facilities do not have access to electricity. For example, about 50%, 39%, 38%, and 31% of healthcare facilities in the Central African Republic, Niger, Chad, and Sierra Leone, respectively, lack access to electricity altogether. When reliability was considered, only 63%, 54%, and 48% of healthcare facilities in Tanzania, Kenya, and Zimbabwe, respectively had reliable access to electricity. This highlights a major infrastructural barrier to the deployment and operation of advanced neuroimaging technologies like MRI that has high power demand and require consistent power supply.

**Table 4:**
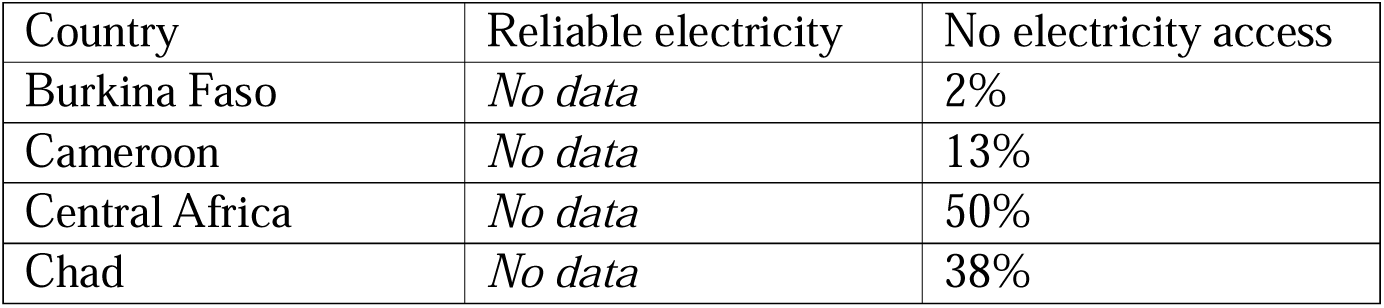
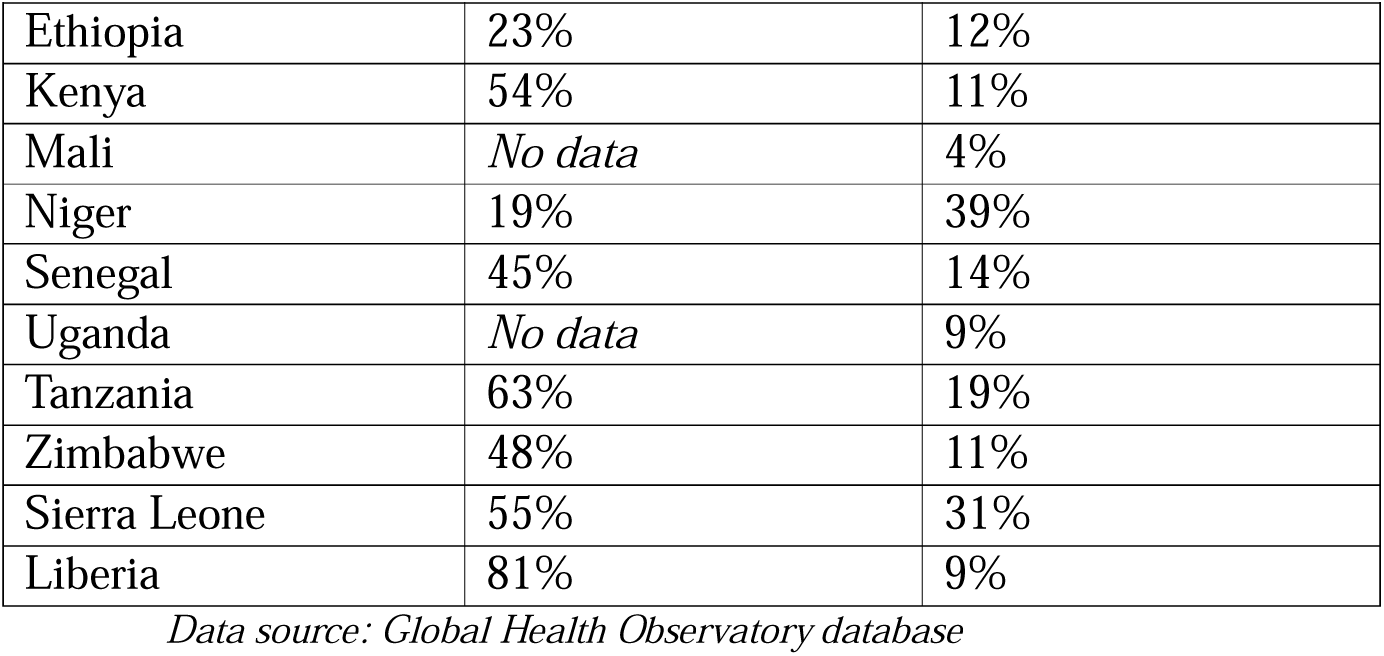
healthcare facilities access to electricity.

| Country | Reliable electricity | No electricity access |
| --- | --- | --- |
| Burkina Faso | *No data* | 2% |
| Cameroon | *No data* | 13% |
| Central Africa | *No data* | 50% |
| Chad | *No data* | 38% |
| Ethiopia | 23% | 12% |
| Kenya | 54% | 11% |
| Mali | *No data* | 4% |
| Niger | 19% | 39% |
| Senegal | 45% | 14% |
| Uganda | *No data* | 9% |
| Tanzania | 63% | 19% |
| Zimbabwe | 48% | 11% |
| Sierra Leone | 55% | 31% |
| Liberia | 81% | 9% |

*Data source: Global Health Observatory database*

### Case study results

Five experts, including radiologists (n =2), a radiology technologist (n =1), a biomedical engineer (n =1), and a nurse (n =1) with experience using or supporting the ULF-MRI were interviewed at Felege Hiwot Comprehensive Specialized Hospital (FCSH). Participants were recruited between March 15, 2026 and April 1, 2026, and written informed consent was obtained from all participants before the interviews.

The researcher also conducted on-site observations at the hospital on the resources needed for - and available to -operate the ULF-MRI, the use of the technology, and the clinical workflow processes. The hospital’s radiology department operates diagnostic technologies such as CT and EEG; however, it does not have a high-field MRI system.

The transition from research into clinical practice

The portable ULF-MRI system was tested and validated for approximately two years at Felege Hiwot Hospital in Ethiopia. During the two-year research phase, the technology was used to assess infants’ brain development, and more than 300 brain scans were conducted. After two years of research, the Hyperfine Swoop® 0.064T MRI system was transitioned into clinical practice, where experts could use it for clinical decision-making.

During the transition of the portable ULF-MRI into clinical practice, several activities, including preventative training, preparation of rooms, and maintenance of the MRI had to be carried out. The MRI engineer of the UNITY project led these activities and the overall transition process. For example, the scanner was operated using its default software version for the duration of the validation research, and it was necessary to update it to the latest software version to benefit from better image quality. Updating the software required up to four hours of uninterrupted power. One of the challenges was frequent power outages at the hospital, which, if it happened during an upgrade, could lead to the malfunction of the ULF-MRI motherboard, as had been reported at other research sites in Pakistan and Bangladesh. If this occurs, the motherboard must be shipped to the US Hyperfine centre for repair. For this reason, a diesel generator had to operate continuously for four hours until the software updating process was completed, resulting in additional fuel costs for the hospital and additional environmental impacts.

Secondly, a group of radiology technologists, biomedical engineers, and a radiologist received three days of in-person preventive maintenance training from the visiting MRI engineer involved in the UNITY project. The purpose of the training was to build local maintenance capacity, enhance troubleshooting skills, and prevent misuse and damage to the machine. The biomedical engineers mentioned that it was their first exposure to an MRI system, but they felt at ease and confident in disassembling and reassembling parts of the ULF-MRI with the guidance of the MRI engineer. One of the biomedical engineers stated:

> *“I have never worked with an MRI machine before. However, I believe that with a bit more training and practice, we will likely be able to perform basic maintenance and replace parts by ourselves. This hands-on training has given me the confidence and foundational knowledge necessary to further enhance my skills as I continue working with the technology.”*

Although the training plan was intended to involve more participants, including physicians, nurses, and other professionals, only a limited number of radiologists and other professionals participated. This was because the hospital is one of the busiest health facilities in the region, dealing with thousands of patients per day, with a limited number of healthcare professionals available. For example, the hospital only has three senior radiologists, and only one of them was able to complete the entire training program due to their demanding work schedules.

Telephone and electrical lines in the room were also removed to avoid potential electromagnetic interference of the imaging process. Finally, radiologists at Felege Hiwot hospital and experts at UNITY together developed an imaging protocol focusing on core MRI sequences like T1-weighted (T1W), T2-weighted (T2W), Fluid-attenuated Inversion Recovery (FLAIR), and Diffusion-Weighted Imaging (DWI). After that, the ULF-MRI was then ready for the clinical brain imaging of both infants and adults. Phantom scanning was performed for quality assurance to calibrate and ensure image quality. They were also recommended to conduct calibration scan regularly.

### Clinical application and acceptability

After completing the training, maintenance, and quality assurance processes, the radiology department began scanning patients who voluntarily agreed to undergo brain imaging for various reasons. Technologists and radiologists who had previously been involved in the two-year research project noted a significant improvement in image quality due to the new software update and the removal of electromagnetic interference in the scanning room.

Reflecting on the image quality one radiologist stated:

> *“I have observed that the image quality has improved with the new software. These images are diagnostically adequate and even have better quality compared to the low-field MRI images patients bring from private clinics.”*

Felege Hiwot Hospital is an early adopter of ULF-MRI technology, but perceptions of its image quality vary among local experts. Some clinicians prefer referring patients to private clinics that have low-field MRI scanners (0.1 and 0.5 T) for brain imaging rather than using the hospital’s ULF-MRI. They already believe that images from low-field MRIs (0.1 and 0.5 T) are of low quality and therefore perceive images from ULF-MRI to be of even lower quality. This perception may be related to the common tendency of equating lower-field MRI with lower clinical utility (1). The UNITY project is currently evaluating and testing the clinical applications of ULF-MRI across multiple sites, which may help address misperceptions regarding its diagnostic capabilities. Additionally, many healthcare professionals at the hospital are not aware of the availability of this technology. Hospital-wide technology awareness and sensitization activities are imperative to address such misperceptions.

### Infrastructure and physical environment needed

During the researcher’s observations of the scanning process, frequent power outages disrupted the scanning process, and scans had to be repeated several times. The hospital did not have an Uninterruptible Power Supply (UPS) to provide backup power for the ULF-MRI or for most of the other equipment in the radiology department and other hospital units. We observed that several dozen images were incomplete or of limited diagnostic value due to power outages and fluctuations during the scanning process. Rather than only coming for a short visit, patients had to stay for hours to repeat the scan as a result. A nurse who was involved in scanning infants mentioned that keeping parents waiting for hours until the power returned was extremely challenging, making it difficult to achieve daily scanning targets. Our study participants recommended either a built-in battery backup for at least 30 minutes or a separate UPS to address the frequent power interruptions and fluctuations.

The Hyperfine Swoop® ULF-MRI requires a stable internet connection to transfer images from the iPad-controlled console to a computer where radiologists interpret the image. The hospital has a dedicated high-speed broadband connection specifically for the ULF-MRI room. However, due to the popularity of this area’s fast internet connection, many hospital staff are also connected to this line, causing the Wi-Fi to be slower for image transfers.

The hospital has allocated a dedicated room with adequate ventilation and the conditions necessary for the safe use of the ULF-MRI scanner. However, electromagnetic interference and noise from the home phone and electrical cables in the room was affecting image quality, requiring the relocation of these cables. The system’s power requirement was compatible with the existing electrical infrastructure of the hospital and worked perfectly with the standard wall outlet.

The researcher observed that most of the hospital’s pathways, corridors, and room entrances are not suitable for moving the scanner from one room to another. Uneven pathways, non-level flooring, and the location of some clinical wards, such as the pediatric ward, on the first floor without elevator access made it difficult to take advantage of the portability feature of the ULF-MRI.

As part of the maintenance and inspection activities conducted during the transition process, the MRI engineer disassembled the ULF-MRI. We found earplugs and rat droppings deep inside the machine, as well as several electrical cables that had been severed by rats and required repair. In addition, regular cleaning of both the room and the machine itself is necessary to address dust issues. Proper cleaning procedures are necessary to ensure the optimal functioning of the equipment, and cleaners must be trained in the correct use of relevant cleaning tools, processes, and materials. At times, cleaners mistakenly used water to clean areas that only need alcohol-based cleaning, although no damage or malfunction has been reported as a result. Windows may also need to have insulation film or protection film to block dust and wind coming through the window frame gaps and cracks.

### Capacity buildingh

As part of the integration of ULF-MRI into the clinical practice of Felege Hiwot Hospital, a brief training session was provided to a small group of participants. However, the participants expressed interest in more advanced training to enhance their technical skills, including integrating AI to improve image quality, and in learning more about the hardware and software details of the ULF-MRI. They believed that these skills’ development could provide independence and create a sustainable imaging ecosystem by enabling local professionals to actively participate in the development and innovation of the technology.

One example cited by the radiologists concerned the new CT scanner the hospital had purchased and its maintenance contract with the vendor. In just a year, the CT scanner required high-level maintenance on more than three occasions. It took months to dispatch maintenance technicians from the vendor in Europe, leaving patients and the hospital without CT imaging services for a total of five months. Experts expressed similar concerns that they may still depend heavily on the ULF-MRI vendor if the scanner requires high-level maintenance. They argued that advanced training programs could reduce such dependency and enhance the long-term sustainability of the technology.

### Human resources and workflow integration

Felege Hiwot hospital has three radiologists and seven radiology technicians serving an extensive catchment area estimated to be more than five million people. Given the number of people this hospital is serving, having only three radiologist consultants could be considered as severe understaffing.

One of the reported advantages of ULF-MRI is its ease of use and reduced reliance on specialized personnel, who are often in short supply in many African countries. During the research and validation phase of the ULF-MRI, nurses operated the scanner, including performing the entire scanning procedures of over 300 infants, with some occasional support from radiology technicians.

The scanner was then transferred to the hospital’s radiology department, where radiology technicians are needed to operate it. Although the intention is for this innovation to be portable within the hospital’s emergency rooms, intensive care units (ICU), and paediatric wards (considering the difficulties mentioned above relating to moving the scanner), it is currently placed in the radiology department. Task shifting from radiologists to other physicians and healthcare professionals is not occurring at present. This may occur at a later stage as the adoption process matures and more professionals outside the radiology department become familiar with the technology. However, radiologists also expressed concerns about relinquishing imaging responsibilities and professional authority to non-radiologists.

One of the radiologists stated:

> *“In my opinion, everyone can operate this technology in our hospital. However, interpreting the images should exclusively remain for radiologists, otherwise our training in this discipline may no longer be necessary quite soon, as innovations in imaging including AI applications are threatening our roles.”*

While some radiologists expressed concerns regarding imaging responsibilities and scope of practice beyond the radiology department, the existing workload suggests that task sharing may become necessary for the efficient use of the technology. This is especially important when considering that the three radiologists and seven radiology technicians were performing hundreds of scans per month using the hospital’s existing imaging systems. The adoption of the ULF-MRI into clinical practice would likely increase the workload and may require hiring more radiology staff unless tasks are shared with other physicians.

We also observed that ULF-MRI is not yet well integrated into the hospital’s imaging workflow. Many hospital departments still had little awareness of this technology. Besides, as mentioned above, some physicians still had concerns about the quality of images the ULF-MRI produces, and they referred patients to private clinics in the city for scans because there is no conventional MRI in the hospital. Three months after the integration of ULF-MRI, there were still no established patient referral pathways or MRI scan requests from other departments.

## Discussion

This study found significant inequities in access to (advanced) neuroimaging services across Africa, driven by not only by the lack of MRI and CT scanners but also by broader weakness of the health system infrastructure. The findings further suggest that portable ULF-MRI may provide a feasible approach to expanding neuroimaging availability in low-resource settings.

As illustrated in this study, the availability of MRI and CT scanners is extremely limited in Africa. Thirty-eight African countries have a combined total of approximately 100 MRI units, compared with 33,000 MRI units across 38 OECD countries. Among the available scanners in Africa, about 39% of them are obsolete but remain in use to meet clinical needs (27). These stark disparities, coupled with ongoing epidemiological and demographic transitions and the growing burden of neurological diseases in Africa (13), are likely to increase the risk of delayed, missed, or incorrect diagnoses of neurological conditions, ultimately adversely affecting health outcomes.

The evidence from this study indicates that the current access gap may not be addressed by simply increasing the number of conventional scanners, as several interconnected constraints remain. Operating these advanced technologies requires multidisciplinary specialized personnel, extensive infrastructure including magnetic and radiofrequency shielding, and reliable high-capacity power supply (12,16,30). However, these resources are in short supply. For example, Malawi and Uganda each have fewer than 10 consultant radiologists, serving populations of 21 and 50 million people, respectively, compared with approximately 5,000 consultant radiologists for 70 million people in the United Kingdom (30,31).

In addition, this study indicated that many healthcare facilities in African countries lack reliable access to electricity. In some countries, such as Central Africa, Chad, Niger, and Sierra Leone, more than one-third of healthcare facilities do not have access to electricity. In line with this, a study in Africa reported that about 57% of MRI facilities experienced downtime at least one to two times per week because of power-related issues (27). These findings suggest that this broader health system weaknesses requires innovative, context-specific, flexible, and affordable solutions.

Given these constraints, portable ULF-MRI systems may help address some accessibility challenges because they are mobile, less expensive, infrastructure-light, and require substantially less specialised expertise (7,30). Our case study of the recent integration of this system into clinical practice in an Ethiopian hospital demonstrated a promising approach in the delivery of neuroimaging in resource-limited settings.

The integration process only required brief hands-on training for the existing staff, and nurses were able to perform scans, thereby reducing dependence on scarce specialists. In addition, the hospital did not require infrastructural modifications, the installation of special radiofrequency shielded room, or a dedicated power supply, as the ULF-MRI integrated well with the existing systems. Similar findings regarding ease of operation, reduced infrastructure requirements, and cost saving have been reported in studies from high and middle income countries (5,12,16,32,33). These characteristics may even enable decentralization of neuroimaging services beyond main hospitals and facilitate access to diagnostic care in semi-urban and rural areas.

Although ULF-MRI overcomes many of the challenges related to conventional MRI and CT, it is observed that the broader health system constraints and many of the infrastructural and operational challenges discussed earlier re-emerged during the clinical integration process in Ethiopia. For example, in Felege Hiwot Hospital, prolonged power outages lasting even several days, as well as frequent short blackouts, are common. These disruptions may affect scan acquisition and potentially damage the MRI scanner itself. The ULF-MRI should therefore incorporate power backup solutions to protect against power outages, surges, and voltage fluctuations. A recent deployment of solar powered ULF-MRI in a rural maternal healthcare centre in Zimbabwe demonstrates the potential alternative and sustainable solution to overcoming the power issues (34).

The integration process at Felege Hiwot Hospital included a brief training on operating the technology and scanning procedures. However, capacity building beyond scanning, particularly in the core engineering and technical aspects of ULF-MRI is essential. Developing and empowering local experts in these core engineering and technical skills will foster sustainability and self-reliance in maintenance, operation, and further innovation (1). This could help healthcare facilities avoid long-term proprietary maintenance contract models typically used in conventional CT and HF-MRI, which are expensive and can exceed the initial cost of the equipment (1).

In addition, to reduce the workload of available radiologists, more extensive training should be provided to non-radiology healthcare practitioners on the nature and characteristics of ULF-MRI images, scanner operations, and safe and effective patient imaging procedures. This would support task shifting, particularly in operating the scanner. Other researchers have also suggested that personnel shortages could be addressed through built-in automated abnormality detection methods, including robust internet connectivity in the design of ULF-MRI to enable tele-radiology support from remote radiologists and also remote connection from technicians (30).

In addition to technical and infrastructural considerations, perception and acceptance of the technology by healthcare professionals are imperative for the successful integration of ULF MRI. A lower magnetic field does not necessarily imply reduced clinical utility. However, some healthcare professionals have such perceptions. The key consideration is whether the images from ULF-MRI have sufficient quality for accurate diagnosis even in a reduced contrast context (1). In line with this, ULF-MRI demonstrated its clinical efficacy in diagnosing various types of neurological conditions including ischemic and haemorrhage stroke, epilepsy, hydrocephalus, multiple sclerosis, and multifocal leukoencephalopathy (16,17,21,35,36). Hospital-wide sensitization workshops and knowledge sharing activities among experts and users may help improve perceptions and address potential misconceptions.

Despite the benefits of ULF-MRI and its promise of improving accessibility, affordability, and flexibility in brain imaging, it still has important technical limitations worth acknowledging. This imaging system has a high degree of noise and artefacts in images, low spatial resolution, and poor signal-to-noise ratio and contrast, which may affect image quality and interpretation (4,12,37,38). Hence, improving image quality using different mechanisms, such as super-resolution technique, the application of artificial intelligence, and deep learning-based reconstruction of images (26,39,40) should be integral to its adoption in standard clinical practice. Furthermore, ULF-MRI is still an evolving technology and needs further validation and testing for many neurological diseases. This limits its current ability to fully replace the roles of HF-MRI in brain imaging, even if ULF-MRI is accessible and affordable.

These findings suggest that efforts to expand neuroimaging services in Africa require not only the availability of diagnostic technologies but also improving the broader healthcare system. Health policymakers and decision-makers may need to invest in workforce development through training and capacity building, as well as in essential infrastructure such as reliable power supply, to sustainably expand access to neuroimaging services. Portable ULF-MRI may complement these efforts by enabling neuroimaging with limited resources and reducing some of the infrastructural barriers often associated with conventional neuroimaging technologies.

## Limitations and future directions

The strength of this study lies in its approach to understand the neuroimaging ecosystem in Africa from a holistic perspective by quantifying availability of MRI and CT scanners, analysing health system challenges, and examining the implementation of portable ULF-MRI into clinical practice in a low-resource setting. However, this study has a few limitations worth noting. The first limitation is the relatively short period between clinical implementation of ULF-MRI and the start of data collection for this study, which was only five months. This short time frame may not provide a complete picture of the extent of utilization, staff opinions, and workflow changes. Future evaluations with a longer follow-up period could provide better insight as the implementation matures.

Secondly, this study was conducted in a regional comprehensive specialized hospital that has a radiology set-up equipped with X-Rays and CT scans. Further implementation studies in other healthcare setting levels including in rural healthcare centres without radiological set-up and tertiary hospitals that already have MRI suites would further generate evidence of integration in diverse clinical setting.

Finally, as health systems vary across countries, similar studies conducted in multiple settings would enhance understanding of contextual, organizational, and systemic-level barriers and facilitators influencing the adoption of ULF-MRI.

Future studies should examine the integration and implementation of portable ULF-MRI in multi-county settings across different health system tiers. It is also important to conduct economic evaluation or cost effectiveness analysis, which will provide evidence for decision makers for wider clinical adoption. In addition, further validation of the technology across a wider range of disease conditions and continued technological improvements to address image quality issues are essential.

## Conclusion

In conclusion, this study showed that the availability of advanced neuroimaging technologies, such as MRI and CT, in Africa remains highly constrained. This limited availability is not only driven by shortage of scanners but also by broader weaknesses in the neuroimaging ecosystem, including a shortage of trained personnel, inadequate health system infrastructure, and affordability issues. ULF-MRI, such as the recently developed 0.064T Hyperfine Swoop® portable system, has the potential to address some of these systemic challenges. The integration of this technology into clinical practice in a hospital in Ethiopia demonstrated the feasibility of deploying ULF-MRI in low-resource settings with limited infrastructural requirements. Capacity building of local experts, particularly in core engineering and technical aspects of the technology, could foster sustainability and self-reliance in maintenance, operation, and further innovation. Further improvements in the technical capacity of the technology, especially in image quality, could enhance its acceptability and scalability. These findings suggest that portable ULF-MRI is a potential pathway to addressing inequalities and expanding neuroimaging access in resource constrained settings.

## Data Availability

All data produced in the present work are contained in the manuscript

## Acknowledgements

N/A

